# Impact of spectrograms on the classification of wheezes and crackles in an educational setting. An interrater study

**DOI:** 10.1101/19005504

**Authors:** JC. Aviles-Solis, I Storvoll, Vanbelle Sophie, H. Melbye

**Author notes:** Corresponding author: Juan Carlos Aviles Solis, UiT The Arctic University of Norway, Post-box 6050 Langnes, 9037 Tromsø.

## Abstract

**Background:** Chest auscultation is a widely used method in the diagnosis of lung diseases. However, the interpretation of lung sounds is a subjective task and disagreements arise. New technological developments like the use of visual representation of sounds through spectrograms could improve the agreement when classifying lung sounds, but this is not yet known.

**Aims:** To test if the use of spectrograms improves the agreement when classifying wheezes and crackles.

**Methods:** We used 30 lung sounds recordings. The sample contained 15 normal recordings and 15 with wheezes or crackles. We produced spectrograms of the recordings. Twenty-three third to fifth-year medical students at UiT the Arctic University of Norway classified the recordings using an online questionnaire. We first showed the students examples of how wheezes and crackles looked in the spectrogram. Then, we played the recordings in a random order two times, first without the spectrogram, then with live spectrograms displayed. We asked them to classify the sounds for the presence of wheezes and crackles. We calculated kappa values for the agreement between each student and the expert classification with and without display of spectrograms and tested for significant improvement. We also calculated Fleiss kappa for the 23 observers with and without the spectrogram.

**Results:** When classifying wheezes 13/23 (1 with p<.05) students had a positive change in k, and 16/23 (2 with p<.05). All the statistically significant changes were in the direction of improved kappa values (.52 - .75). Fleiss kappa values were k=.51 and k=.56 (p=.63) for wheezes without and with spectrograms. For crackles, these values were k=.22 and k=.40 (p=<0.01) in the same order.

**Conclusions:** The use of spectrograms had a positive impact on the inter-rater agreement and the agreement with experts. We observed a higher improvement in the classification of crackles compared to wheezes.

## Introduction

Chest auscultation is a widely used method in the diagnosis and follow up of several diseases. Medical doctors use it to guide clinical decisions and treatment strategies. (1) However, the identification and interpretation of the sounds remains a subjective task. Generally, auscultation occurs in a “solitary” fashion since normal stethoscopes are designed for individual listening. Often, disagreement between health professionals arises when classifying lung sounds and the reliability in the classification of wheezes and crackles has been found to be moderate at best. (2-6) This variation is caused by not only diverse identification, but also due to different labeling of the sounds. (7, 8) These limitations of auscultation make the training of new health professionals a challenging task. It has been suggested that difficulties in teaching and learning auscultation also contribute to the demise of this technique. (9)

However, new electronic stethoscopes can capture and store sounds in a digital form. Assisted by the processing capacity of personal computers and/or mobile phones it is possible to generate visual representations of sound in the form of spectrograms. In spectrograms, the common adventitious lung sounds, wheezes and crackles, show recognizable patterns, (figure 1) which may be of help in the identification of these sounds. (10) Some of the electronic stethoscopes available on the market offer this solution. The use of spectrograms also gives the possibility for group analysis and discussion. In addition, the use of recordings with spectrograms could avoid the objectification of patients used in the teaching of auscultation. (11)

**Figure 1.**
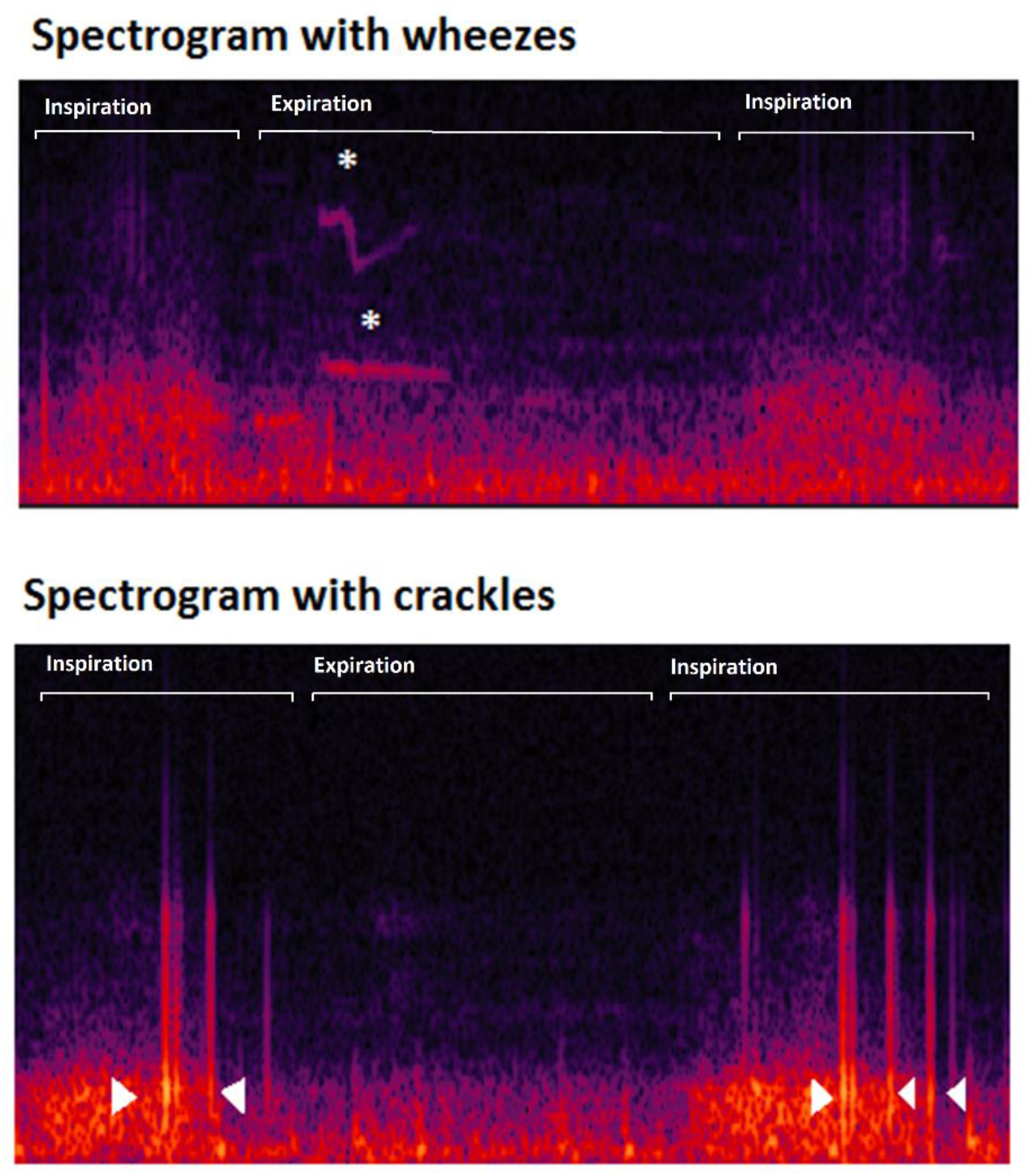
Examples of spectrograms of lung sound recordings showing the presence of wheezes (stars) and crackles (arrowheads.

It would be natural to think that the use of a visual support could help to improve the classification of lung sounds since there are two sensory inputs instead of one. The use of spectrograms could improve the teaching of lung sounds by giving an aid to the listening abilities in training. Andrés et al (12) found that the spectrograms do have a positive impact on how medical students assign a diagnosis with the help of lung sounds. However, the design of this study did not isolate the effect of spectrograms and therefore this affirmation is inconclusive. There are very few studies on this subject. Therefore, we do not know whether the use of spectrograms would help to medical students to better classify lung sounds. If spectrograms have a positive effect in the classification of lung sounds it would make it a valuable tool to make auscultation training more simple and effective.

The aim of this study is to explore how the use of spectrograms affects the agreement between medical students and a panel of experts and the agreement within a group of students in classifying lung sounds.

## Methods

### Data for classification

We recorded lung sounds from 20 adults. They were 67.4 years old on average (44–84) and nine were female. We registered the following information about the subjects: age, gender and self-reported history of heart or lung disease. No personal information was registered that could link the sound recordings to the individual subjects. The project was presented for the Regional Committee for Medical and Health Research Ethics, and it was considered to be outside the remit of the Act on Medical and Health Research.

To record the lung sounds, we used a microphone MKE 2-EW with a wireless system EW 112-P G3-G (Sennheiser electronic, Wedemark, Germany) placed in the tube of a Littmann Master Classic II stethoscope (3M, Maplewood, MN, USA) at a distance of 10 cm from the headpiece. The microphone was connected to a digital sound Handy recorder H4n (Zoom, Tokyo, Japan).

We placed the membrane of the stethoscope against the naked thorax of the subjects. We asked the subjects to breathe deeply while keeping their mouth open. We started the recording with an inspiration and continued for approximately 20 seconds trying to capture three full respiratory cycles with good quality sound. We performed this same procedure at six different locations (figure 2). The researcher recording sounds used a headphone as an audio monitor to evaluate the quality. When too much noise or cough was heard during the recording, a second attempt was performed.

**Figure 2.**
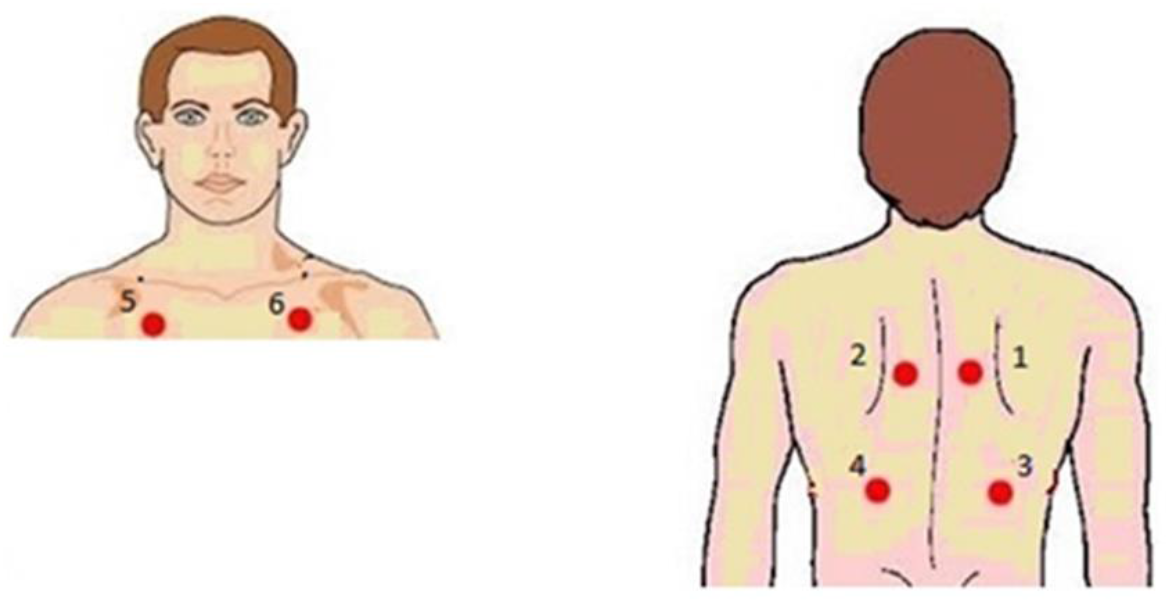
Illustration showing the different places where lung sounds were recorded. (1_2) Between the spine and the medial border of the scapula at the level of T4–T5; (3_4) at the middle point between the spine and the mid-axillary line at the level of T9–T10; (5_6) at the intersection of the mid-clavicular line and second intercostal space.

We obtained 120 audio files in ‘.wav’ format and recorded at a sample rate of 44 100 Hz and 16 bit depth in a single monophonic channel. We did not perform post-processing of the sound files or implement filters. We chose 30 recordings for this study from 18 different subjects. This selection contained 15 normal and 15 abnormal sounds, from which nine were classified as containing crackles and six containing wheezes.

A panel of four experts in the field of lung sound research classified the recordings according to the presence of wheezes and crackles by a majority criterion.

### Observers

We invited medical students from third to fifth year at UiT, The Arctic University of Norway. The students had received the standard curricular training in physical examination provided in the medicine school. The training in lung auscultation included a lecture on lung sounds which included the demonstration of some spectrograms. Beyond this, the students had no previous experience with the use of spectrograms.

### Presentation and classification of the lung sounds

First, we presented a couple of recordings with wheezes and crackles and showed the students how these sounds looked in the spectrograms. Then we played the 30 recordings in a random order in two sessions. In the first session, we presented sound only. In the second session, the sound and the spectrograms were simultaneously displayed in the classroom screen. There was a pause of 20 minutes in between the two sessions. We presented no additional information beyond the sound and the spectrograms. The observers were not aware that the same sounds were played in both sections.

In both sessions, each recording was played two times and the students had up to 30 seconds to classify it before moving to the next recording. The observers used their personal computers and an online classification scheme (Questback AS, Norway). In this scheme, the observers had to specify if the recording contained only normal respiratory sounds. If this was not the case, the observers had to further specify if the recording contained wheezes, crackles or other sounds and if they appeared during inspiration or expiration. It was also possible to mark the recording as containing too much noise to be classified. At the end of the classifications session we obtained a report in an excel document. (Microsoft,Redmond,WA,USA)

### Statistical analysis

We calculated Cohen kappa for the agreement between the observers and the experts, and Fleiss kappa for all the observers as a group. We then compared the kappa values obtained in the sections with and without the use of spectrograms and calculated p values to explore for statistically significant differences using an adaption of Hotelling’s T2 test described by Vanbelle, S. (13) In this analysis, the recordings were clustered by the individual they were recorded from. We used Holm’s correction procedure to adjust p values for multiple hypothesis testing. We used R version 3.2.1 and the package “magree” to perform all the calculations. Significance level was set at p <0.05. In addition, we calculated sensitivity and specificity of each participant using the experts’ classification as the gold standard. We tested for significant differences in sensitivity and specificity with and without the intervention using paired Wilcoxon signed rank test with continuity correction.

The results of this study are reported according to the Guidelines for Reporting Reliability and Agreement Studies (GRRAS). (14)

## Results

### Observers

We included 23 observers in the study. At the beginning, 30 students accepted to participate in the study. From them, two withdrew before the start of the study, two did not show up and three did not complete the classification session due to lack of time. Eight participants were third year students, fourteen participants were from the fourth year and one was from the fifth year. There were 19 women and four men.

### Agreement

The students observed a mean prevalence of wheezes of 9.7 (6 – 15) without spectrograms and 8.3 (5 – 12) with spectrograms. In the case of crackles the students observed a mean prevalence of 11.5 (4 – 22) and 10.9 (5 – 18) in the same order. The mean proportion of agreement (%) and Cohen kappa (k) with the experts for all 23 participants classifying wheezes without spectrograms was 82 % and k=.56. We observed 88 % and k=.68 with the use of spectrograms. In the case of crackles we observed a proportion of agreement of 72 % and k=.38 without spectrograms. With the use of spectrograms 80 % and k=.56. Fleiss kappa values for the multirater agreement were k=.51 and k=.56 (p=.63) for wheezes without and with spectrogram, respectively. For crackles, we observed k=.22 and k=.40 (p=<0.01) in the same order. (figure 3) Compared to the expert panel’s classification, 13/23 students had a positive change in kappa when classifying wheezes (one with p<.05), and 16/23 (two with p<.05) when classifying crackles. (figure 4 and figure 5) All the statistically significant changes were in the direction of improved kappa values (.52 - .75).

**Figure 3.**
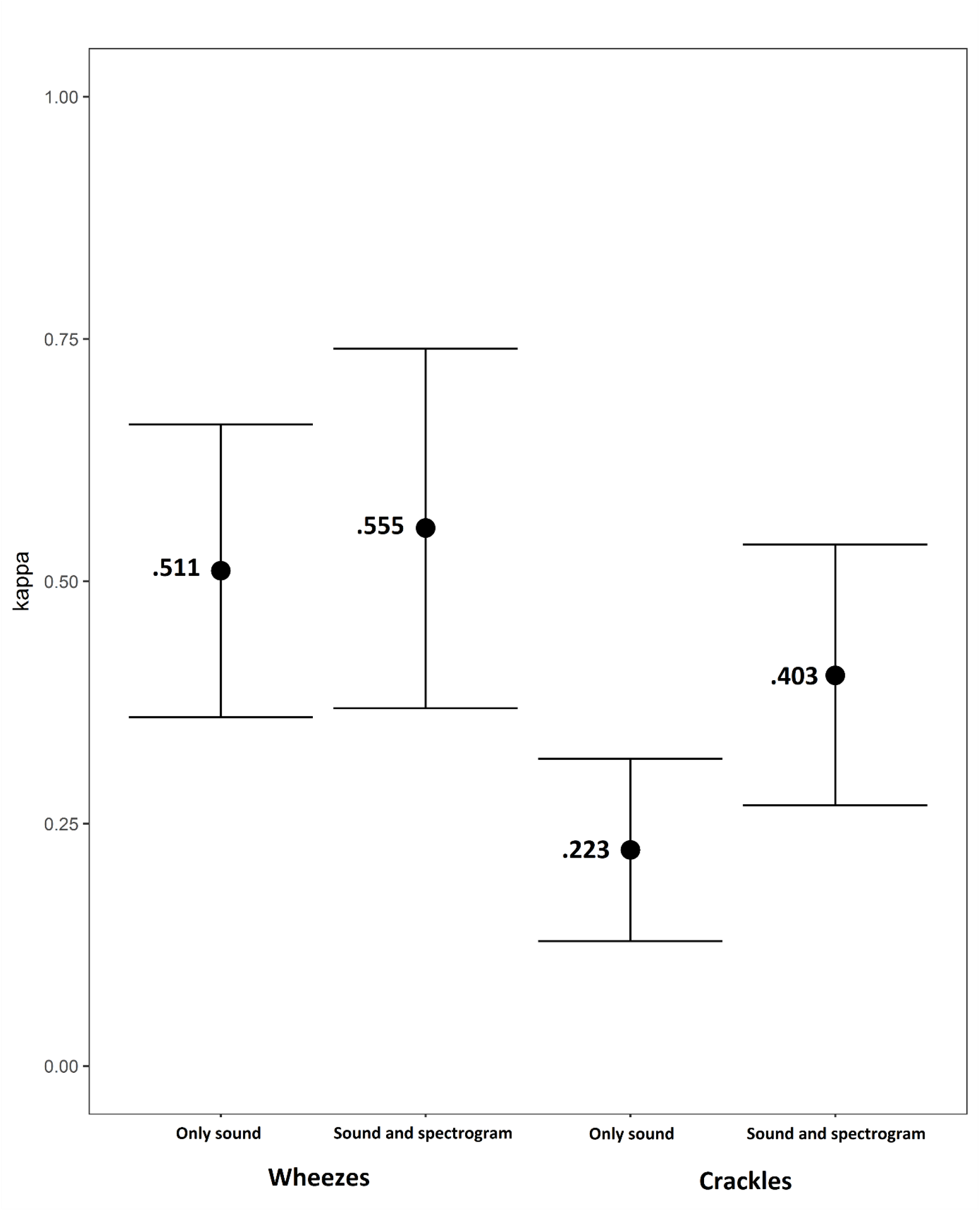
Fleiss kappa for the group of 23 participants when classifying wheezes and crackles with only sound and sound plus spectrogram.

**Figure 4.**
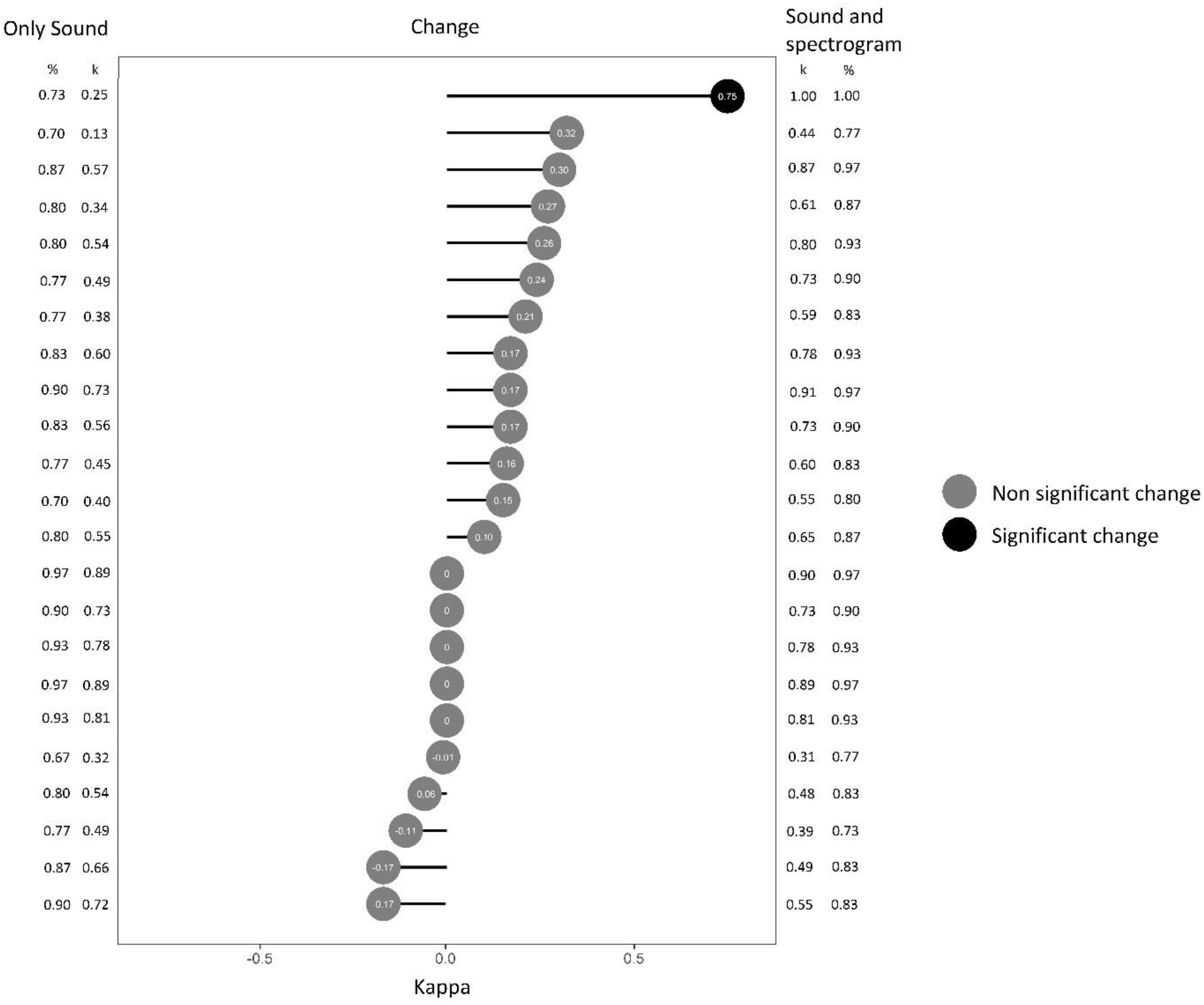
Cohen’s kappa of each participant when classifying wheezes with only sound (left) and with sound and spectrograms (right) compared to the reference standard. The change in kappa between the two classifications and its statistical significance is illustrated at the center. Proportion of agreement (%) is presented on the lateral columns.

**Figure 5.**
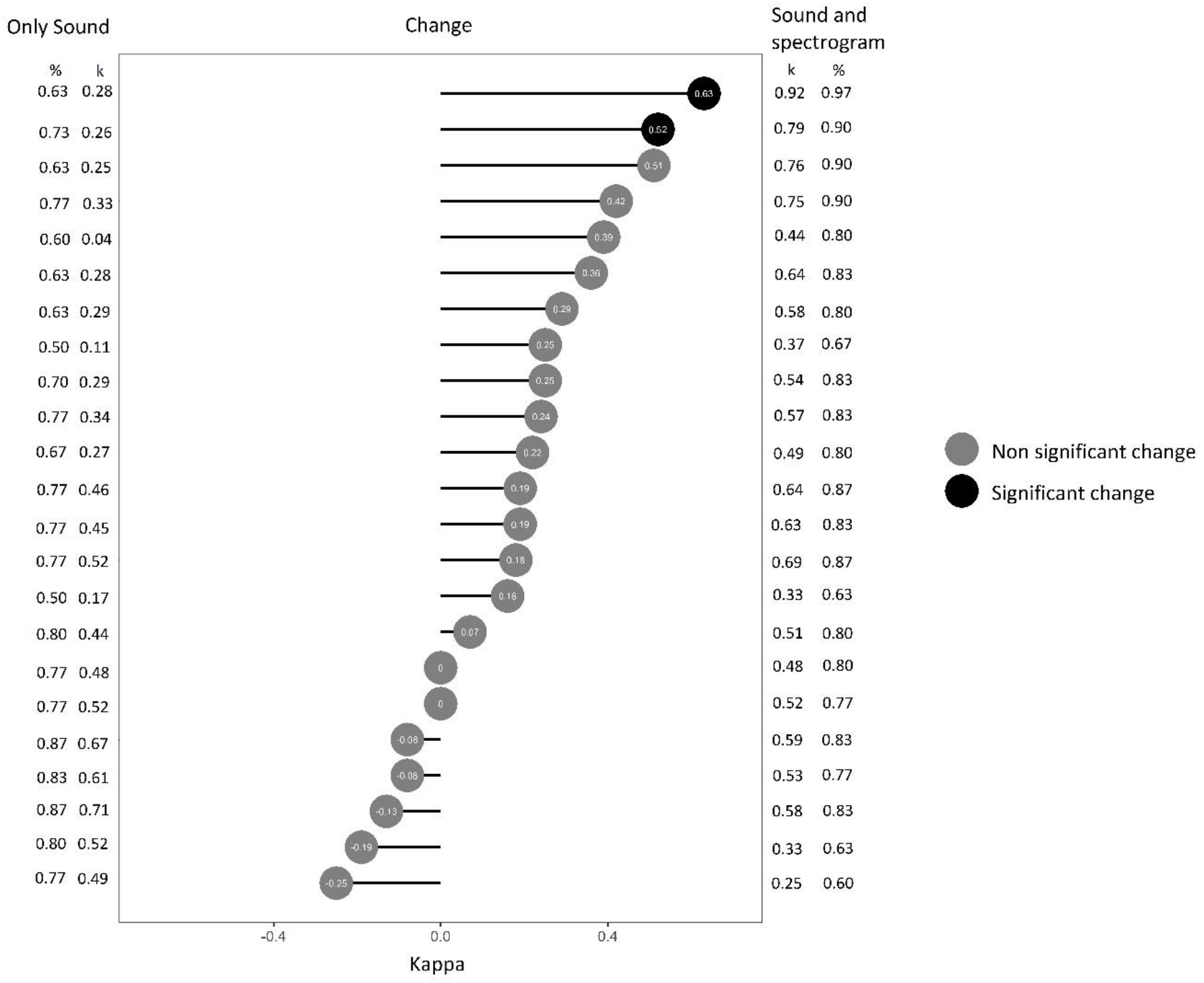
Cohen’s kappa of each participant when classifying crackles with only sound (left) and with sound and spectrograms (right) compared to the reference standard. The change in kappa between the two classifications and its statistical significance is illustrated at the center. Proportion of agreement (%) is presented on the lateral columns.

When looking at the classification of normal vs abnormal sounds (wheezes or crackles) we observed a mean prevalence of abnormal sounds of 18.7 and 18 with and without spectrogram (experts 15). The mean absolute agreement was 72 % with a mean kappa of k= 0.44 without spectrograms and 80% and k=0.60 with spectrograms. Only one participant had a significant improvement in this analysis.

The median sensitivity for wheezes did not present a significant change but the specificity was higher in the classification with the use of spectrograms (p=0.002). In the case of crackles, there was a significant increase in sensitivity (p=0.03) when using spectrograms but without significant change in specificity. (figure 6)

**Figure 6.**
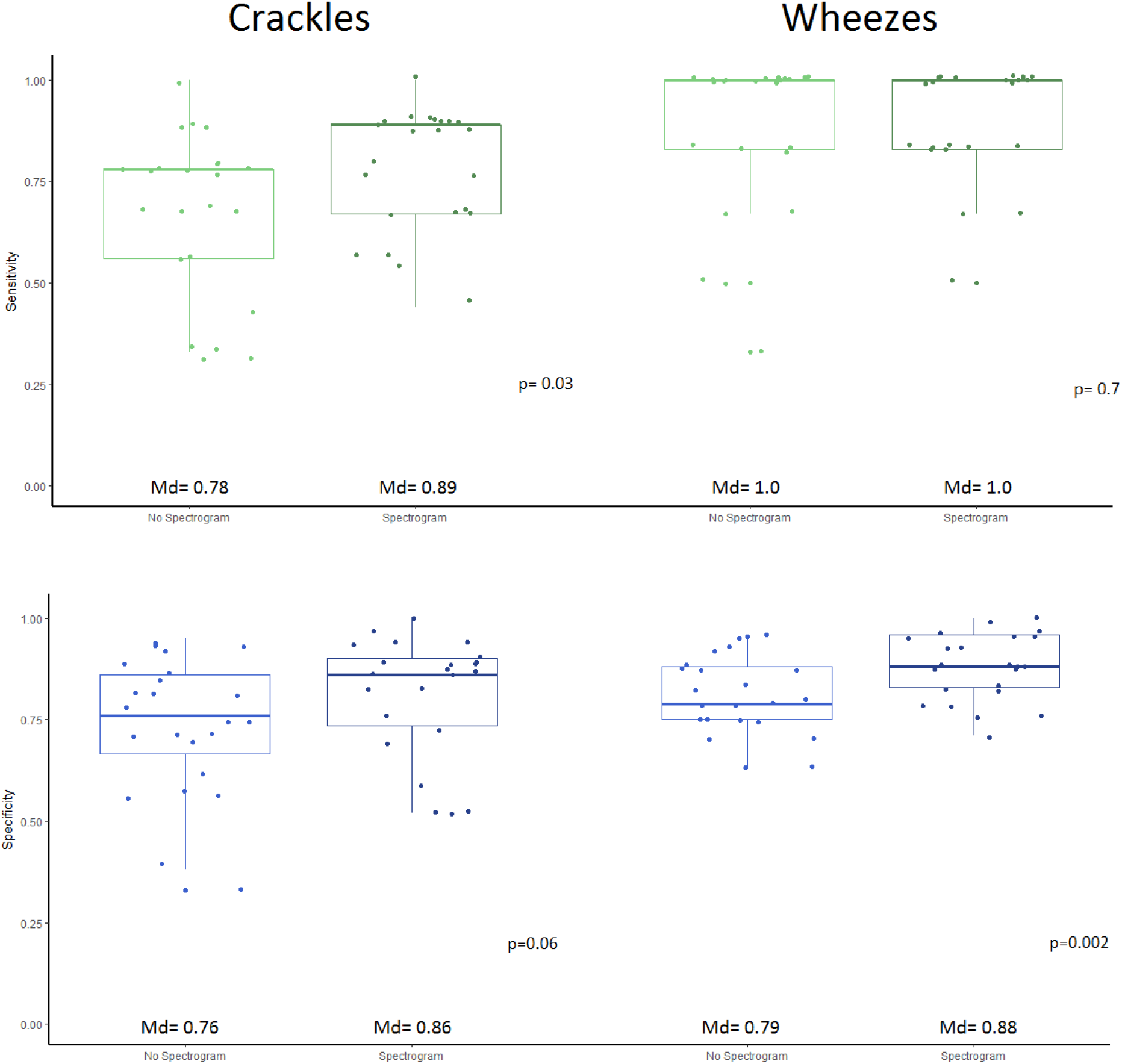
Box and whiskers diagrams showing the change in sensitivity and specificity of the students classifying wheezes and crackles with and without spectrograms. The answers of the experts were considered as the reference standard. P values shown were obtained from the test of difference between means using paired Wilcoxon signed rank test with continuity correction. Md= Median.

## Discussion

We found improved agreement with the experts in the classification of lung sounds with the use of spectrograms. However, most of the improvements were not statistically significant. We did observe a significant improvement in the agreement within the group (Fleiss kappa) when classifying for crackles when the sounds were presented with spectrograms.

The levels of individual and group agreement observed in this study corresponds with that reported in previous studies. (15) We generally observed a higher agreement for wheezes than for crackles, in accordance with what is described in the literature. (3) It is interesting that the impact of spectrograms was different for wheezes and crackles. It might be that wheezes are easier to recognize without spectrograms due to its relatively long duration and its musical quality, which make them more familiar to the human ear. On the contrary, crackles are short explosive sounds that could easily be missed by ear appreciation or perceived as noise. (16) For this reason, having a visual aid could be an advantage to identify them.

Andrés et al (12) found that the spectrograms do have a positive impact on how medical students assign a diagnosis to a patient with the help of lung sounds. Even though the observer populations are similar, the results are not comparable since the outcome in their test was a diagnosis and not the classification of the sounds. Since the participants in Andrés’ study got clinical information together with the sound this could have influenced how they classified lung sounds. Nguyen et al observed that the addition of clinical information has an effect on the classification of lung sounds and this effect is experience-dependent. In Nguyen’s study, the group classifying lung sounds without clinical information achieved similar scores classifying lung sounds regardless of clinical experience. When clinical information was provided, then the more experienced raters achieved higher scores. (17)

Our exploratory study suggests that the use of spectrograms might be helpful to improve the teaching of auscultation, mostly in the cases of crackles. This by enhancing interrater agreement and facilitating discussion of the sounds in a teaching arena.

### Strengths and limitations

We think the methodology employed allowed us to analyze the isolated effect of the spectrograms in the classifications.

The students had free seating. Even though absolute silence was required during the classification, we cannot fully rule out the possibility that the students could have influenced each other. This could have influenced the estimation of agreement in the group (multi-rater) agreement in a positive direction.

We have tested the same hypothesis 23 times. This increases the chance of making a type I error. We have taken this situation into account by correcting the p-values with Holm’s procedure.

Some limitations in our study concern the sample size of sounds to classify. Due to the exploratory nature of this study, we chose a number of sounds to be classified which could allow us to perform the whole procedure in a time window of two hours. This, to avoid dropouts from the study. It is possible that we could have had observed more significant changes if the sound sample would have been larger. Future studies looking at the effect of spectrograms in agreement should take this into account in its design.

## Conclusion

The use of spectrograms had a positive impact on the inter-rater agreement and the agreement compared to experts. We observed a higher improvement in the classification of crackles compared to wheezes.

## Data Availability

Data can be available upon reasonable request to the corresponding author.

## Acknowledgements

We would like to thank Professor A. Marques, Professor P. Piirilä, Professor H. Pasterkamp and Professor A. Sovijarvi for their help in classifying the lung sound recordings used in this study. We would like to thank Dr. Cristina Jácome for its valuable insight. The authors also would like to thank LHL rehabilitation centre at Skibotn, Norway, for its cooperation in performing the recordings.

